# Researcher and Clinician Preferences for a Journal Transparency Tool: A Mixed-Methods Survey and Focus Group Study

**DOI:** 10.1101/2024.05.28.24307345

**Authors:** Jeremy Y. Ng, Henry Liu, Mehvish Masood, Jassimar Kochhar, David Moher, Alan Ehrlich, Alfonso Iorio, Kelly D. Cobey

## Abstract

**Background:** Transparency within biomedical research is essential for research integrity, credibility, and reproducibility. To increase adherence to optimal scientific practices and enhance transparency, we propose the creation of a journal transparency tool (JTT) that will allow users to obtain information about a given scholarly journal’s operations and transparency policies. This study is part of a program of research to obtain user preferences to inform the proposed JTT. Here, we report on our consultation with clinicians and researchers.

**Methods:** This mixed-methods study was conducted in two parts. The first part involved a cross-sectional survey conducted on a random sample of authors from biomedical journals. The survey asked clinicians and researchers about the inclusion of a series of potential scholarly metrics and user features in the proposed JTT. Quantitative survey items were summarized with descriptive statistics. Thematic content analysis was employed to analyze text-based responses. Subsequent focus groups used the survey responses to further explore the inclusion of items in the JTT. Items with less than 70% agreement were used to structure discussion points during these sessions. Participants voted on the use of user features and metrics to be considered within the journal tool after each discussion. Thematic content analysis was conducted on interview transcripts to identify the core themes discussed.

**Results:** A total of 632 participants (5.5% response rate) took part in the survey. A collective total of 74.7% of respondents found it either ‘occasionally, ‘often’, or ‘almost always’ difficult to determine if health information online is based on reliable research evidence.

Twenty-two participants took part in the focus groups. Three user features and five journal tool metrics were major discussion points during these sessions. Thematic analysis of interview transcripts resulted in six themes. The use of registration was the only item to not meet the 70% threshold after both the survey and focus groups. Participants demonstrated low scholarly communication literacy when discussing tool metric suggestions.

**Conclusions:** Our findings suggest that the JTT would be valuable for both researchers and clinicians. The outcomes of this research will contribute to developing and refining the tool in accordance with researchers and clinicians.

## Background

The outcomes of biomedical research are most commonly communicated through publications in scholarly journals. Maintaining research integrity (e.g., requiring ethical approval, peer review, plagiarism checks, and indexing in authorized databases) and the quality [1] of publications through transparency and open practices is essential for clinical and research decision-making [2,3]. However, concerns that some journals operate in a ‘black box’ and are not forthcoming about their processes have been raised [4]. For example, in an editorial published in *Science* [4], it was argued that science would be improved if journals permitted and participated in empirical research and quality assurance of their peer review processes. Challenges also exist surrounding how clinicians and researchers seek information to inform their decision-making and writing. Despite Google and Google Scholar placing no quality controls on what is indexed [5], studies have found that 60–90% of clinicians use Google to find information to help them make point-of-care choices [6–8] and many researchers routinely search Google Scholar in conjunction with, and sometimes even as a replacement for other bibliographic databases, when conducting systematic reviews [9,10].

Further, journals that have been reported to engage in suboptimal transparency practices and flawed peer-review processes have started to infiltrate legitimate archiving systems such as PubMed/MEDLINE [11]. As a result of such challenges, clinicians and researchers require a mechanism to discern journal quality.

To address these concerns and increase transparency practices among scholarly journals, we propose developing an automated journal transparency tool (JTT) that users could employ to obtain information about a given scholarly journal’s operations and transparency policies [12]. As we envision it, users (e.g., researchers, clinicians, patients) could then use this data to make a more informed decision about whether or not they want to engage with the journal (e.g., read it, submit manuscripts to it, or cite work published there). Here, we obtained preferences for the proposed JTT from the clinician and researcher communities. This tool is part of a wider initiative whereby we are using a user-centered design strategy [13,14], to obtain stakeholder preference for patients [15] and publishers [16].

To collect researchers’ and clinicians’ preferences for the proposed JTT, we conducted a mixed-methods study in two parts: a survey and a focus group. The study is descriptive, and we have no a priori hypotheses. The researcher/clinician communities’ views on what should be included in a JTT will contribute to meaningfully situating it within the scholarly landscape and help to ensure that the most relevant inputs are used to build the tool.

## Methods

### Research Ethics Approval and Transparency Practices

Research ethics board approval for the study was obtained from the Ottawa Health Science Network Research Ethics Board (REB ID # 20230041-01H). The study protocol was registered on the Open Science Framework (OSF) [17] and can be found at https://doi.org/10.17605/OSF.IO/6EWQS [18]. The online survey used within the first part of the study was informed by items in the checklist for reporting results of internet e-surveys (CHERRIES) reporting guidelines [19] and the focus groups conducted within the second part of the study were informed by the consolidated criteria for reporting qualitative research (COREQ) checklist [20]. Individual participant data from the survey was anonymous, while individual participant data from the focus groups was anonymized; data that was shared publicly using OSF was anonymous or deidentified.

### Study Design

This study consisted of two parts: a cross-sectional survey and a focus group.

### Part 1: Survey

We designed a purpose-built survey containing questions relating to: (1) demographic characteristics (5 items); (2) practices associated with published research literature (7 items); (3) user feature preferences (i.e., how the JTT user interface should look, how data automation can facilitate the metrics the JTT reports, and how to disseminate a completed JTT to the community and track its uptake; 4 items); and (4) JTT metrics (i.e., metrics that users will access about each *individual* journal on the JTT to make informed decisions regarding the use of that journal for clinical or research purposes; 17 items). This survey, which contained both quantitative and qualitative (free-text) questions, was piloted by a group of researchers and clinicians who were not part of the study. The survey was created and administered via the University of Ottawa’s approved version of SurveyMonkey [21]. For the complete survey, please see https://osf.io/7nu24.

#### Identifying Participants

Similar to an approach used in previously published studies [22–24], a convenience sample of 12 000 random authors who published articles no earlier than June 2022 within biomedical journals on MEDLINE was selected for researcher and clinician recruitment. To be eligible, participants needed to be an author/co-author of a biomedical article and be able to read and write in English. A standardized recruitment script that invited researchers and clinicians to participate in our survey was created and emailed to participants. Invitees received an initial email on April 24, 2023, followed by three reminder emails, each spaced 1 week apart. The final email was sent on May 15, 2023, and the survey closed on May 25, 2023. All participants were presented with an implied informed consent form (see https://osf.io/f479j) prior to being able to see the survey questions and were required to confirm that they gave their consent to participate prior to beginning the survey.

#### Analysis of Survey Data

We report the overall response for each quantitative item, as well as descriptive statistics such as frequencies and percentages. Thematic content analysis was used to identify, analyze, and report patterns or ’themes’ within qualitative text-based items [25]. All responses to each open-ended question were reviewed and coded inductively [26] by two researchers independently (HL, JK). Members of the research team (HL, JK, MM) discussed themes iteratively until all the themes and subthemes for each question were identified and agreed upon.

### Part 2: Focus Groups

Survey participants were invited to a follow-up online focus group that was structured using the Nominal Group Technique [27] (5-7 participants per group, 3-6 groups to aim to identify approximately 90% of themes [28]). Focus groups were conducted between October 30, 2023, and November 10, 2023, and were approximately 1 hour long each. Prior to the start of the first focus group involving survey participants, the focus group was piloted by JYN, HL, KDC, DM, and two other researchers who were not involved in the design/conduct of this study. All survey participants who agreed to participate were provided with a consent form and gave their consent verbally prior to taking part in a session. We developed a discussion guide informed by the results of the survey, where survey items with less than 70% agreement as the main discussion points for the focus groups. This 70% agreement threshold was selected based on past literature [29]. A mock prototype of the JTT was presented, and four steps were followed based on the Nominal group technique [27]:

- ‘Silent generation of ideas’ where participants brainstormed individually
- ‘Round robin’ where participants each share one of their ideas, one at a time, until there are no new responses
- ‘Discussion’ where participants refine ideas together
- ‘Voting and ranking’ where participants will rank their preferred items.

JYN, who was a postdoctoral fellow at the time of the study, conducted the focus groups. He explained the purpose of the study and the discussion process, acted as a moderator, and sought permission to audio/video record the sessions. A separate research member (HL) took field notes during the session. Both researchers had no prior relationship with participants, and no personal information about the researchers (e.g., biases, assumptions) was disclosed to focus group participants. Prior to beginning the focus group, verbal consent was obtained from all participants. Sessions were video and audio recorded using Zoom software. No one other than the participants and researchers were present during the focus groups. Demographic information was not collected from participants, and transcripts were not returned to participants for review.

#### Analysis of Focus Group Data

Automated interview transcripts, which were reviewed for accuracy by members of the research team, were used to conduct a thematic content analysis to derive themes from the data [25]. First, focus group notes were combined, uploaded, and inductively coded [30] into Microsoft Excel independently by two researchers (HL, MM). Following that, two research members (HL, MM) met to compare codes and evaluate them for inclusion based on whether they directly addressed the discussion point, and then iteratively discussed themes until themes and subthemes were established. The lead author (JYN) had training in qualitative interviewing and provided training and supervision to HL and MM. Data saturation was not discussed, and participants did not provide feedback on the findings.

## Results

### Part 1: Survey

A total of 632 participants responded to the survey, representing a response rate of 5.5% (632/11,554). The survey achieved a completion rate of 75%, with respondents having an average completion time of 9 minutes 29 seconds. Participant demographics are summarized in **Table 1**. Quantitative items (**Table 2**) were first examined, followed by qualitative survey results (**Table 3**). Aggregate survey responses (https://osf.io/wfvp6) and survey analysis data (https://osf.io/3p5kf) have been made available on OSF.

**Table 1:** Survey Participant Demographic Characteristics.

| Characteristic | Response (n, %) | N |
| --- | --- | --- |
| Gender | Man: 361 (59.0)<br>Woman: 247 (40.4)<br>Other: 4 (0.7) | 612 |
| Age | 18-29 yrs: 39 (6.3)<br>30-39 yrs: 200 (32.4)<br>40-49 yrs: 193 (31.3)<br>50-59 yrs: 121 (19.6)<br>60+ yrs: 64 (10.4) | 617 |
| Top 5 Countries of Work | United States of America: 134 (22.1)<br>Canada: 36 (5.9)<br>China: 30 (4.9)<br>Australia: 30 (4.9)<br>Italy: 27 (4.4) | 607 |
| Research/Clinician Status<br>(Check all that apply) | Undergraduate Student: 3 (0.5)<br>Graduate Student: 45 (7.3)<br>Postdoctoral Fellow: 61 (10)<br>Research Coordinator/Associate/Analyst: 35 (5.7)<br>Clinician: 112 (18.3)<br>Assistant Professor: 101 (16.5)<br>Associate Professor: 143 (23.3)<br>Professor: 138 (22.5)<br>Researcher affiliated with Industry: 20 (3.3)<br>Research affiliated with Government: 40 (6.5)<br>Other: 46 (7.5) | - |
| Career Stage | Early career (<5 yrs): 124 (20.2)<br>Mid-career (5-10 yrs): 183 (29.8)<br>Senior career (>10 yrs): 307 (50) | 614 |

**Table 2:**
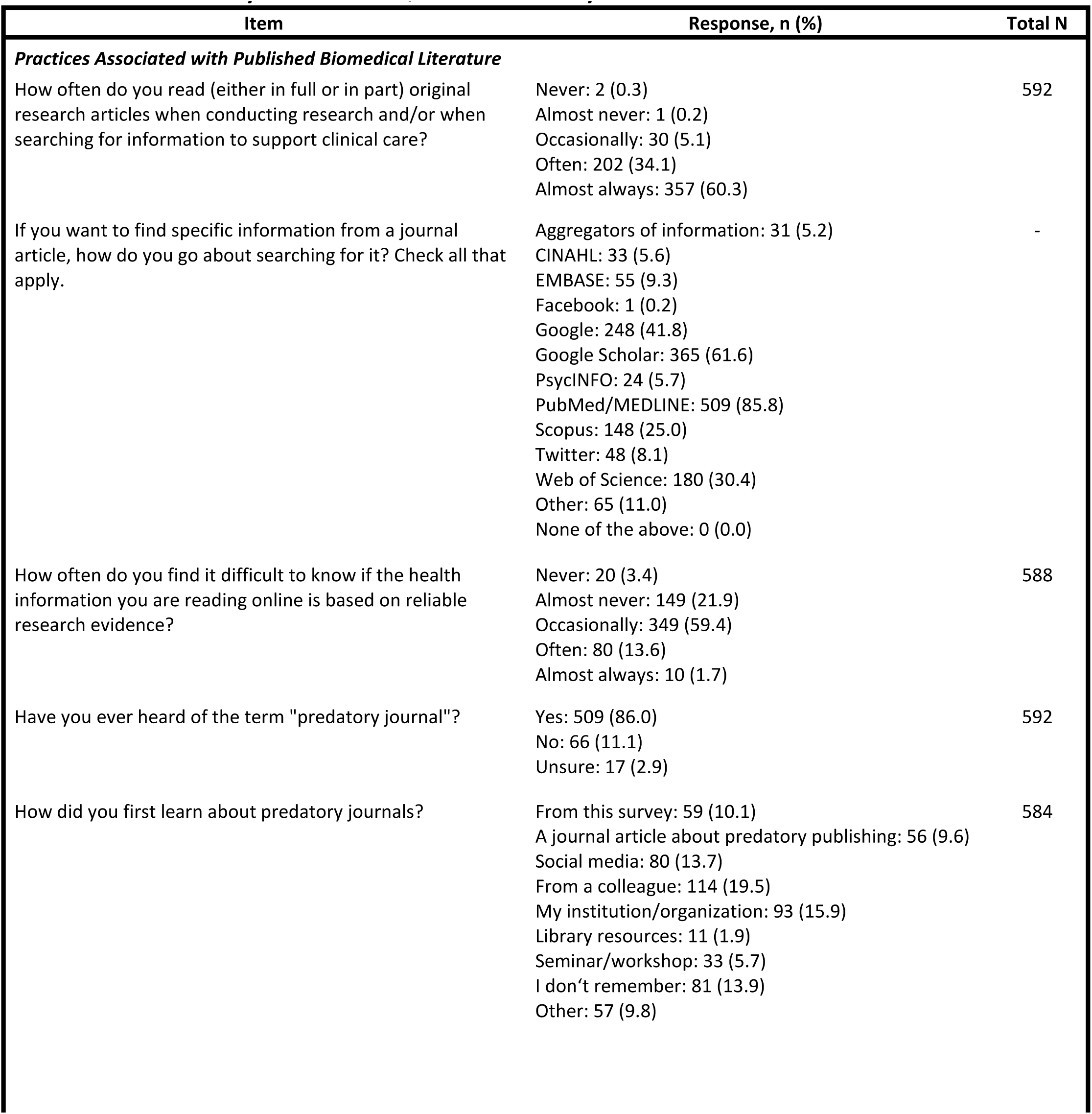
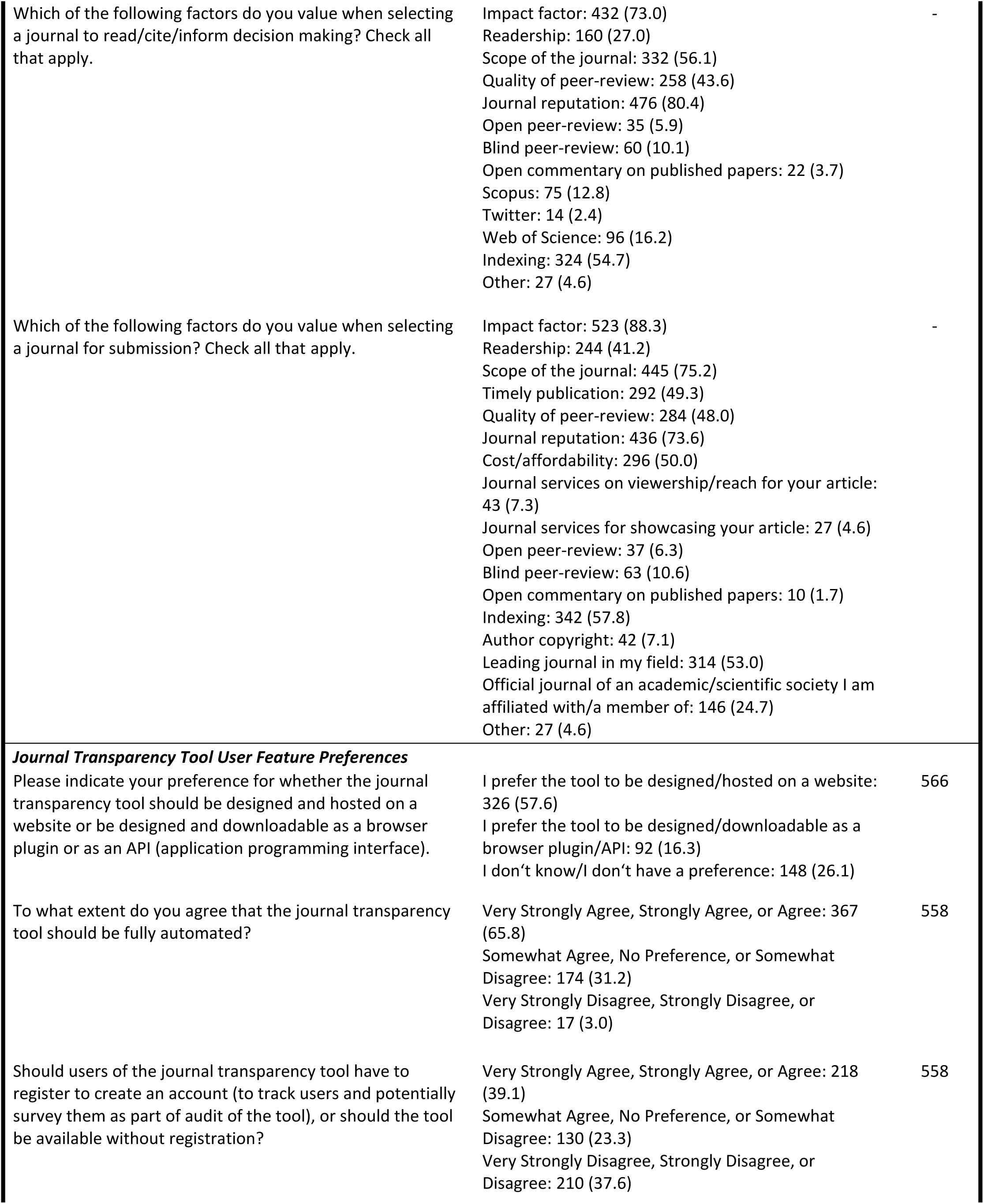
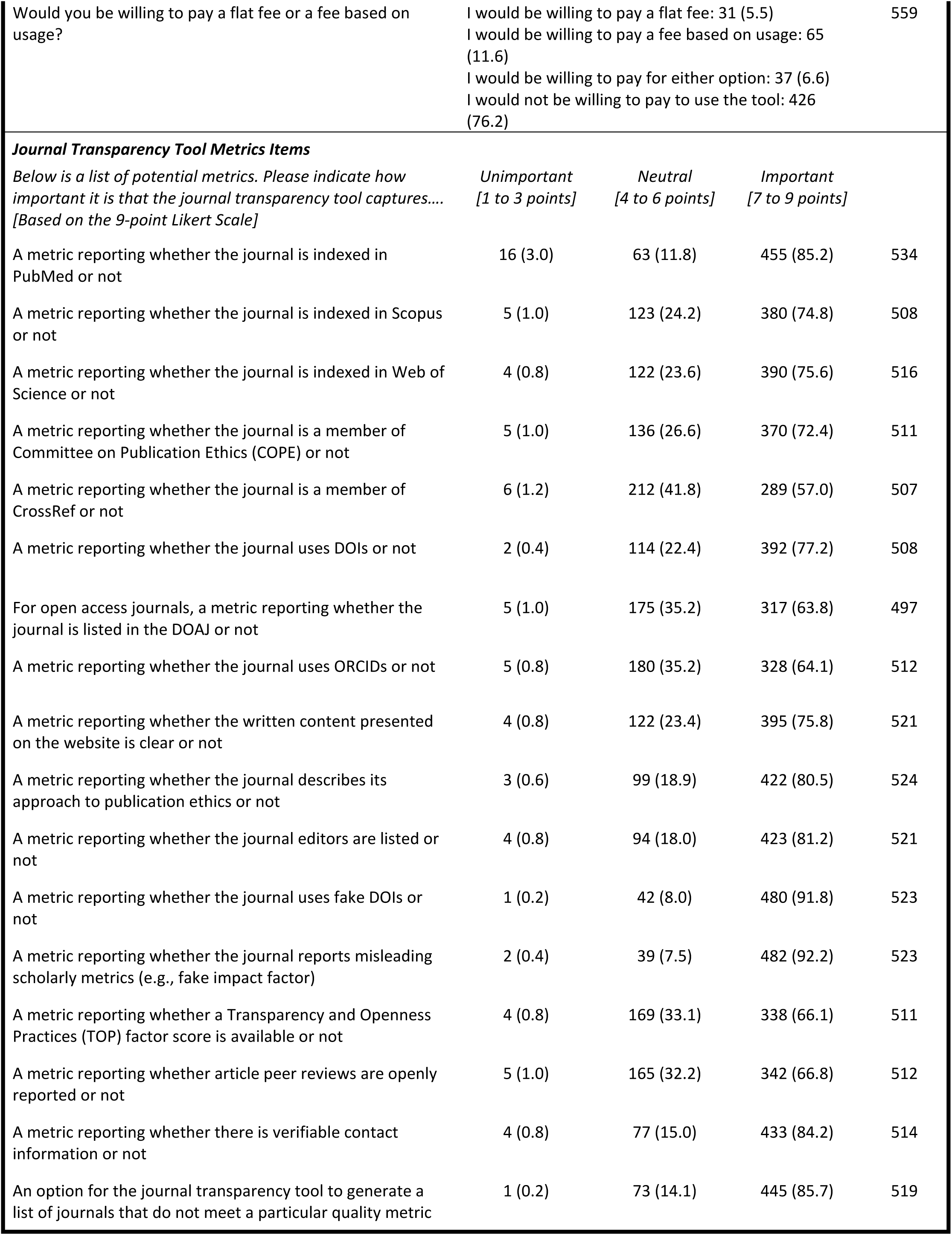
Summary of Results of Quantitative Survey Items.

| Item | Response, n (%) | Total N |
| --- | --- | --- |
| ***Practices Associated with Published Biomedical Literature*** |  |  |
| How often do you read (either in full or in part) original research articles when conducting research and/or when searching for information to support clinical care? | Never: 2 (0.3)<br>Almost never: 1 (0.2)<br>Occasionally: 30 (5.1)<br>Often: 202 (34.1)<br>Almost always: 357 (60.3) | 592 |
| If you want to find specific information from a journal article, how do you go about searching for it? Check all that apply. | Aggregators of information: 31 (5.2)<br>CINAHL: 33 (5.6)<br>EMBASE: 55 (9.3)<br>Facebook: 1 (0.2)<br>Google: 248 (41.8)<br>Google Scholar: 365 (61.6)<br>PsycINFO: 24 (5.7)<br>PubMed/MEDLINE: 509 (85.8)<br>Scopus: 148 (25.0)<br>Twitter: 48 (8.1)<br>Web of Science: 180 (30.4)<br>Other: 65 (11.0)<br>None of the above: 0 (0.0) | - |
| How often do you find it difficult to know if the health information you are reading online is based on reliable research evidence? | Never: 20 (3.4)<br>Almost never: 149 (21.9)<br>Occasionally: 349 (59.4)<br>Often: 80 (13.6)<br>Almost always: 10 (1.7) | 588 |
| Have you ever heard of the term "predatory journal"? | Yes: 509 (86.0)<br>No: 66 (11.1)<br>Unsure: 17 (2.9) | 592 |
| How did you first learn about predatory journals? | From this survey: 59 (10.1)<br>A journal article about predatory publishing: 56 (9.6)<br>Social media: 80 (13.7)<br>From a colleague: 114 (19.5)<br>My institution/organization: 93 (15.9)<br>Library resources: 11 (1.9)<br>Seminar/workshop: 33 (5.7)<br>I don't remember: 81 (13.9)<br>Other: 57 (9.8) | 584 |
| Which of the following factors do you value when selecting a journal to read/cite/inform decision making? Check all that apply. | Impact factor: 432 (73.0)<br>Readership: 160 (27.0)<br>Scope of the journal: 332 (56.1)<br>Quality of peer-review: 258 (43.6)<br>Journal reputation: 476 (80.4)<br>Open peer-review: 35 (5.9)<br>Blind peer-review: 60 (10.1)<br>Open commentary on published papers: 22 (3.7)<br>Scopus: 75 (12.8)<br>Twitter: 14 (2.4)<br>Web of Science: 96 (16.2)<br>Indexing: 324 (54.7)<br>Other: 27 (4.6) | - |
| Which of the following factors do you value when selecting a journal for submission? Check all that apply. | Impact factor: 523 (88.3)<br>Readership: 244 (41.2)<br>Scope of the journal: 445 (75.2)<br>Timely publication: 292 (49.3)<br>Quality of peer-review: 284 (48.0)<br>Journal reputation: 436 (73.6)<br>Cost/affordability: 296 (50.0)<br>Journal services on viewership/reach for your article: 43 (7.3)<br>Journal services for showcasing your article: 27 (4.6)<br>Open peer-review: 37 (6.3)<br>Blind peer-review: 63 (10.6)<br>Open commentary on published papers: 10 (1.7)<br>Indexing: 342 (57.8)<br>Author copyright: 42 (7.1)<br>Leading journal in my field: 314 (53.0)<br>Official journal of an academic/scientific society I am affiliated with/a member of: 146 (24.7)<br>Other: 27 (4.6) | - |
| **Journal Transparency Tool User Feature Preferences** |  |  |
| Please indicate your preference for whether the journal transparency tool should be designed and hosted on a website or be designed and downloadable as a browser plugin or as an API (application programming interface). | I prefer the tool to be designed/hosted on a website: 326 (57.6)<br>I prefer the tool to be designed/downloadable as a browser plugin/API: 92 (16.3)<br>I don't know/I don't have a preference: 148 (26.1) | 566 |
| To what extent do you agree that the journal transparency tool should be fully automated? | Very Strongly Agree, Strongly Agree, or Agree: 367 (65.8)<br>Somewhat Agree, No Preference, or Somewhat Disagree: 174 (31.2)<br>Very Strongly Disagree, Strongly Disagree, or Disagree: 17 (3.0) | 558 |
| Should users of the journal transparency tool have to register to create an account (to track users and potentially survey them as part of audit of the tool), or should the tool be available without registration? | Very Strongly Agree, Strongly Agree, or Agree: 218 (39.1)<br>Somewhat Agree, No Preference, or Somewhat Disagree: 130 (23.3)<br>Very Strongly Disagree, Strongly Disagree, or Disagree: 210 (37.6) | 558 |

|  |  |  |  |  |
| --- | --- | --- | --- | --- |
| Would you be willing to pay a flat fee or a fee based on usage? | I would be willing to pay a flat fee: 31 (5.5)<br>I would be willing to pay a fee based on usage: 65 (11.6)<br>I would be willing to pay for either option: 37 (6.6)<br>I would not be willing to pay to use the tool: 426 (76.2) |  |  | 559 |
| **Journal Transparency Tool Metrics Items** |  |  |  |  |
| *Below is a list of potential metrics. Please indicate how important it is that the journal transparency tool captures....*<br>*[Based on the 9-point Likert Scale]* | *Unimportant*<br>*[1 to 3 points]* | *Neutral*<br>*[4 to 6 points]* | *Important*<br>*[7 to 9 points]* |  |
| A metric reporting whether the journal is indexed in PubMed or not | 16 (3.0) | 63 (11.8) | 455 (85.2) | 534 |
| A metric reporting whether the journal is indexed in Scopus or not | 5 (1.0) | 123 (24.2) | 380 (74.8) | 508 |
| A metric reporting whether the journal is indexed in Web of Science or not | 4 (0.8) | 122 (23.6) | 390 (75.6) | 516 |
| A metric reporting whether the journal is a member of Committee on Publication Ethics (COPE) or not | 5 (1.0) | 136 (26.6) | 370 (72.4) | 511 |
| A metric reporting whether the journal is a member of CrossRef or not | 6 (1.2) | 212 (41.8) | 289 (57.0) | 507 |
| A metric reporting whether the journal uses DOIs or not | 2 (0.4) | 114 (22.4) | 392 (77.2) | 508 |
| For open access journals, a metric reporting whether the journal is listed in the DOAJ or not | 5 (1.0) | 175 (35.2) | 317 (63.8) | 497 |
| A metric reporting whether the journal uses ORCIDs or not | 5 (0.8) | 180 (35.2) | 328 (64.1) | 512 |
| A metric reporting whether the written content presented on the website is clear or not | 4 (0.8) | 122 (23.4) | 395 (75.8) | 521 |
| A metric reporting whether the journal describes its approach to publication ethics or not | 3 (0.6) | 99 (18.9) | 422 (80.5) | 524 |
| A metric reporting whether the journal editors are listed or not | 4 (0.8) | 94 (18.0) | 423 (81.2) | 521 |
| A metric reporting whether the journal uses fake DOIs or not | 1 (0.2) | 42 (8.0) | 480 (91.8) | 523 |
| A metric reporting whether the journal reports misleading scholarly metrics (e.g., fake impact factor) | 2 (0.4) | 39 (7.5) | 482 (92.2) | 523 |
| A metric reporting whether a Transparency and Openness Practices (TOP) factor score is available or not | 4 (0.8) | 169 (33.1) | 338 (66.1) | 511 |
| A metric reporting whether article peer reviews are openly reported or not | 5 (1.0) | 165 (32.2) | 342 (66.8) | 512 |
| A metric reporting whether there is verifiable contact information or not | 4 (0.8) | 77 (15.0) | 433 (84.2) | 514 |
| An option for the journal transparency tool to generate a list of journals that do not meet a particular quality metric | 1 (0.2) | 73 (14.1) | 445 (85.7) | 519 |

**Table 3:** Summary of Results of Qualitative Survey Item Responses.

| Item | Theme (Subthemes) |
| --- | --- |
| Please indicate your preference for whether the journal transparency tool should be designed and hosted on a website or be designed and downloadable as a browser plugin or as an API.<br>With respect to the last question, do you have any other comments to share on the above options or alternatives? | **Factors to Consider in Tool Format** (Tool Availability, Ease of Use, Linking with Other Platforms, Data Security Concerns)<br>**Support for Website** (Plugin/API Performance Issues, Popular Website Integration, Website Increases Accessibility, Bookmarks, Limited Familiarity with API Technology)<br>**Support for API** (Increasing Future Research, Recurring Visits) |
| To what extent do you agree that the journal transparency tool should be fully automated?<br>With respect to the last question, please provide the rationale for your choice. | **Support for Automation** (Humans Are Error-prone, Automated Tool Increases Objectivity, Automation is More Resource Efficient)<br>**Opposition for Automation** (Human Knowledge is Superior, Automation Can Produce Incorrect Outputs, Journal Exclusion, Unable to Measure Metrics, Automation Reduces Objectivity, Humans Correcting Tool Errors)<br>**Automation Suggestions** (Stakeholder Feedback, Automated Journal tool Description) |
| The journal transparency tool will be made publicly available. Should users of the journal transparency tool have to register to create an account, or should the tool be available without registration?<br>With respect to the last question, please provide the rationale for your choice. | **Support for Registration** (Tool Enhancement, Tracking Tool Misuse, Optional Registration, External Platform Integration, Registration Journal Tool Description)<br>**Opposition for Registration** (Data Safety Concerns, Registration is a Barrier to Entry) |
| Would you be willing to pay a flat-fee or a fee based on usage?<br>Please share any other general comments regarding the journal transparency tool user design. | **Opposition to Tool Fee** (Conflicts of Interest, Reduced Global User Access, Rising Costs, Junior Researchers)<br>**Suggestions for Tool Fee Structure** (Flat Fee Structure, Alternative Funding Sources, Opposed Funding Sources, Usage-based Fee Structure, Additional Paid Features) |
| Are there any metrics not listed within the survey that you think would be valuable to include? | **Journal Characteristics** (General Journal Features, Journal History, Turnaround Time)<br>**Journal Guidelines** (Submission Guidelines/Policies, Publication Fees/Funding Models, Open Science, Peer Review Policy/Practices)<br>**Article/Journal Reach** (Alternative Metrics, Traditional Metrics, Readership Metrics)<br>**Equity and Diversity** (Equity and Diversity)<br>**Academic Misconduct and Suspect Practices** (Conflicts of Interest, Fraud and Plagiarism, Editorial Ethics/Legitimacy, Retraction/Withdrawal, Journal Predatory Nature) |
| Please share any other comments regarding the proposed information the journal transparency tool will measure and present. | **Journal Credibility and Accuracy** (Conflicts of Interest, Legitimate Journal Exclusion, Data Availability and Trustworthiness)<br>**Equity and Inclusion** (Equity-Deserving Groups, User Involvement)<br>**Journal Tool Format and Metrics** (Tool Format and Design, Journal Policies and Standards Metrics, Journal Reach Metrics, Peer Review Metrics, Article Publication Metrics) |

#### Demographic Characteristics

Most respondents were male (n = 361, 59.0%) and a plurality were 30 to 39 years old (n = 200, 32.4%). Participants worked in 78 countries worldwide, with individuals from the United States having the highest representation (n = 134, 22.1%). Respondents came from a variety of research and clinical-based backgrounds, with associate professors (n = 143, 23.3%) having the highest representation of participants and undergraduate students (n = 3, 0.5%) having the lowest. Fifty percent (n = 317) of participants had more than 10 years of work experience.

#### Practices Associated with Published Biomedical Literature

Most participants indicated that they ‘often’ (n = 202, 34.1%) or ‘almost always’ (n = 357, 60.3%) read original research articles when conducting research or when searching for information regarding clinical care. PubMed/MEDLINE (n = 509, 85.8%) and Google Scholar (n = 365, 61.6%) were the most used sources to find information. The top four factors for choosing a journal to read, cite, or inform decision-making and for selecting a journal when submitting articles for publication were the same according to respondents: journal reputation, impact factor, scope of journal, and indexing. Most participants ‘almost never’ (n = 149, 21.9%), ‘occasionally’ (n = 349, 59.4%), and ‘often’ (n = 80, 13.6%) found it difficult to know if health information online is based on reliable research evidence and had previously heard of the term predatory journal (n = 509, 86.0%). Ten percent (n = 59) of participants heard of predatory journals for the first time from this survey.

#### JTT User Feature Preferences

The only journal tool preference category that met the general agreement threshold of 70% was fee payment structure. Seventy-six percent (n = 426) of participants indicated that they would not be willing to pay a fee to use the tool. The use of registration was contentious, with 39.1% (n = 218) indicating that it should be necessary, 37.6% (n = 210) disagreeing with its use, and 23.3% (n = 130) being neutral. Many participants (n = 326, 57.6%) indicated that they would prefer that the tool be hosted on a website, while a smaller proportion (n = 92, 16.3%) preferred a browser plugin/API (application programming interface) format. Fully automating the tool was supported by 65.8% (n = 367) of participants.

#### JTT Metric Items

Every journal metric item suggested for the tool was rated as either ‘important’ or ‘neutral’ by most participants on the 9-point Likert scale [34]. Only a small percentage, ranging from 0.2% to 3.0%, regarded any individual metric as ‘unimportant’. The use of fake DOI (digital object identifiers; n = 480, 91.8%) and whether the journal reports misleading scholarly metrics (e.g., fake impact factors; n = 482, 92.2%) were the two metrics that the highest portion of participants identified as ‘important’. Out of the 17 suggested metrics, five (CrossRef, Directory of Open Access Journals [DOAJ], Open Researcher and Contributor ID [ORCID], TOP factor, and open peer review practices) did not reach the 70% threshold of general agreement. A large proportion of participants, ranging from 32.3% to 41.8%, were ‘neutral’ to the use of these five items specifically.

#### Qualitative Survey Responses

Eighteen themes and 64 subthemes were created from six survey items (see **Table 3**). Participants provided reasons to support and oppose the implementation of several JTT features (e.g., the use of registration, website vs. browser plugin/API interface formats, fully automating the tool) within multiple qualitative responses. Similar to quantitative survey feedback, respondents generally did not express support for the use of a tool fee in their answers, but some did give suggestions for the tool fee structure. More than 70 additional journal metrics that were not part of the survey were suggested by participants. When participants were asked about any other comments that they had concerning the JTT, responses tended to focus on journal credibility and accuracy; equity and inclusion; and journal tool format and metrics.

### Part 2: Focus Groups

A total of 22 participants took part in five focus groups. Each focus group had three to seven participants. Within the survey, three items related to user feature preferences (the use of registration, website or browser plugin/API format, and full tool automation) and five journal tool metric suggestions (CrossRef, DOAJ, ORCID, TOP factor, and open peer review practices) did not reach the 70% general agreement threshold. All eight items were major discussion points during the focus groups, where participants provided their thoughts on each item and were subsequently required to vote on their preferences for the incorporation of each item into the tool (see **Table 4** for voting results). Analysis of focus group interview transcripts resulted in a total of six themes (see **Table 5** for thematic summary). Focus group analysis data (https://osf.io/6x7jg) have been made available on OSF.

**Table 4:** Focus Group Voting Results.

| Item | Responses (n, %) |  |  | Total N |
| --- | --- | --- | --- | --- |
|  | Yes | No | Abstain |  |
| ***Journal Transparency Tool User Feature Preferences*** |  |  |  |  |
| Should users of the journal transparency tool have to register to create an account? | 6 (28.6) | 13 (61.9) | 2 (9.5) | 21 |
| Should the Journal Transparency Tool should be fully automated? | 16 (80.0) | 3 (15.0) | 1 (5.0) | 20 |
| Should the Journal Transparency Tool be designed and hosted on a website, as opposed to a browser plugin? | 19 (90.5) | 1 (4.8) | 1 (4.8) | 21 |
| ***Journal Transparency Tool Metrics Items*** |  |  |  |  |
| *Should the Journal Transparency Tool include...* |  |  |  |  |
| A metric reporting whether a journal is a member of CrossRef? | 15 (78.9) | 2 (10.5) | 2 (10.5) | 19 |
| A metric reporting whether the journal is listed in the DOAJ? | 18 (94.7) | 2 (5.3) | 0 (0.0) | 19 |
| A metric reporting whether the journal uses ORCID? | 14 (77.8) | 2 (11.1) | 2 (11.1) | 18 |
| A metric reporting whether a Transparency and Openness Practices (TOP) factor score is available? | 17 (100.0) | 0 (0.0) | 0 (0.0) | 17 |
| A metric reporting whether article peer reviews are openly reported? | 16 (94.1) | 0 (0.0) | 1 (5.9) | 17 |

**Table 5:** Focus Group Thematic Analysis.

| Theme | Subtheme | Codes | Samples Quotes <sup>a</sup> |
| --- | --- | --- | --- |
| Registration | Support for registration | Tracking Tool Impact<br>Part of the Process | "... tracking the impact..." (P16)<br>"...seems part of the process." (P1) |
|  | Concerns with registration | Barrier to Entry<br>Passwords<br>Time Consuming<br>Privacy<br>No value added | "...a potential barrier to entry" (P21)<br>"...a source of frustration " (P11)<br>" .... a little cumbersome for the sake of time" (P8)<br>" ...concerned about...people accessing my data." (P2)<br>"...doesn't seem like I get a lot of value." (P2) |
|  | Suggestions and solutions for registration | Two account types<br>Increase Value<br>Additional Features | "...two type of accounts, one that does not this registration and another one" (P3)<br>"... added value for registering, then I'd feel, perhaps more incentive to do so" (P14)<br>" ...linking it with ORCID" (P20) |
| Full Automation | Support for full automation | Increased Feasibility<br>Resource Constraints<br>Autonomy from Journals<br>Encourage Journal Transparency | "...increases feasibility" (P21)<br>"...budget constraints only allow...fully automated" (P11)<br>".... wouldn't trust if journals are giving in their own input" (P8)<br>".... [encourages] journals commit to display...most transparent" (P18) |
|  | Concerns with full automation | Checking for Accuracy<br>Automation not possible for all metrics | "...have someone by checking [the information]" (P10)<br>"...tough to say everything we want could be possible from full automation." (P1) |
| Website vs. Plugin | Both formats have different purposes | Different functions<br>Target difference audiences | "...depend on the use of the tool" (P5)<br>"...differ [with]... their target population users" (P13) |
|  | Websites are easier to access | Websites are accessible<br>Plugins are difficult to install<br>Plugin technological issues<br>Plugin privacy concerns | "...website is very simple to go to." (P19)<br>"...not a lot of flexibility to be able to put plugins on our browsers" (P9)<br>"...might be causing some glitches" (P8)<br>" ...don't know what... will be collected." (P14) |
|  | Both formats are easy to navigate | Websites are easier to use<br>Plugins are easier to use | "...more easy way to look for the ISBN number" (P10)<br>"...click on the plugin and then just use it there" (P8) |
| Journal Metric Items (CrossRef, DOAJ, ORCID, TOP Factor) | Mixed opinions on reporting items | Items are important<br>Items are not essential<br>Importance relative to individual | "... [these] metrics are quite important and critical" (P10)<br>"...don't really see the value" (P1)<br>"We all exist in our own little bubble." (P21) |
|  | Low item literacy | Not familiar with items<br>Open to learning | "...first time I've understood this material." (P7)<br>"...have to know a little bit more... [but] like the idea" (P1) |
| Open Peer Review (OPR) | General support for reporting OPR | • Support for OPR<br>• Not familiar with OPR | "...nothing against including it" (P19)<br>"...not totally sure about the pros and cons" (P16) |
| Journal tool descriptions | Tool component clarity | • Registration purpose<br>• Use of full automation | "if the point of registration is solely to track users... making that like very explicit" (P21)<br>"...defined in that way that the tool provides...fully automated information" (P11) |
|  | Increase literacy | • Journal tool items descriptions<br>• Open peer review practices descriptions | "...having those descriptions... [that] this is what these metrics encompass" (P21)<br>"...[to know] if the journal has a double or triple blind revision" (P3) |

<sup>a</sup> P refers to participant ID.

#### Registration

Focus group participants had mixed views regarding registration. Reasons to support registration included tracking the tool’s impact and registration being a normal part of the process. Concerns with registration included it being a barrier to entry, time-consuming, appearing to add no value, requiring the use of passwords, and a potential invasion of privacy. During focus group voting, registration was the only category that did not meet the 70% agreement threshold, with 28.6% (n = 6) of participants supporting its use, 61.9% (n = 13) not providing support, and 9.5% (n = 2) abstaining. Implementing two types of accounts (i.e., one where users do not need to register and another where users register to access specialized features), increasing the value for registration, and adding additional features (e.g., linking the tool to ORCID) were mentioned as ways to increase support for registration.

#### Full Automation

A total of 80% (n = 16) of participants voted for the use of full automation following the focus group discussion. Reasons provided to support full automation included increased feasibility, resource constraints, encouraging journal transparency, and autonomy from journals/publishers. Concerns with automation included the necessity for humans to check for accuracy and that automation is not possible for all metrics.

#### Website vs. Plugin

Participants acknowledged that the tool format depends on the purpose of the tool because website and browser plugins have different functions and target different audiences. Websites were viewed as easier to access, primarily because plugins may be more difficult to install, many have experienced plugin technological issues, and participants had data privacy concerns regarding how much and what type of information plugins could collect. Both formats were considered to be easy to navigate when finding information. Ninety percent (n = 19) of participants supported the use of the website during focus group voting.

#### Journal Metric Items

Many participants presented with confusion and expressed a lack of previous knowledge when discussing journal metric items (i.e., CrossRef, DOAJ, ORCID, and TOP Factor). Participants requiring additional information about each journal metric were generally unopposed to their use. Stronger opinions were presented by participants who had previous experience with the journal metrics. The value of each metric was acknowledged to be relative to individual experience, including but not limited to career level and country of work.

#### Open Peer Review

No participant appeared to oppose the use of open peer review as a metric during focus group discussions and voting. However, none of the participants clearly articulated why having such a metric was valuable. Some participants acknowledged that they were not familiar with the idea of open peer review practices. Many provided personal opinions on the use of open peer review in general without specifically elaborating on why it may be a valuable metric to add to the JTT.

#### Journal Tool Descriptions

Participants across all eight major discussion topics stated that providing clear descriptions should be a part of the tool. Descriptions were described as necessary for explaining the purpose of various components of the tool (e.g., the purpose of registration and what collected data from the registration process would be used for). Additionally, descriptions were viewed as essential to increasing the literacy of users who may not have had prior experience with journal tool metrics (e.g., CrossRef) so that users may be able to make informed decisions when choosing a journal.

## Discussion

This two-stage study allowed us to obtain clinician and researcher preferences for a proposed JTT. We first gained information regarding researcher and clinician preferences through a survey. Then, we conducted focus groups to further explore survey responses. Obtaining participant practices related to published biomedical literature allowed us to evaluate the usefulness of such a tool for researchers and clinicians. The two most common resources used to find journals were PubMed/MEDLINE (n = 509, 85.8%) and Google Scholar (n = 365, 61.6%). Previous studies have noted that journals engaging in suboptimal transparency practices have started to infiltrate both archiving systems [11,31]. Furthermore, most respondents (n = 439; 74.7%) found it either ‘occasionally, ‘often’, or ‘almost always’ difficult to determine if health information online is based on reliable research evidence, and approximately 10% (n = 59) of participants had not previously heard of predatory journals prior to this survey. These results are consistent with prior research indicating that a considerable number of individuals encounter challenges in assessing the reliability of online information [32,33]. Additional resources, such as the JTT that we are proposing, are likely to be necessary to increase adherence to transparency and open practices.

Low scholarly communication literacy presented by participants further supports the need for this tool. Many focus group respondents were unfamiliar with scholarly journal metrics (e.g., CrossRef, DOAJ, and TOP factor). Further, although participants had prior knowledge of open peer review practices, they were unable to clearly delineate the advantages and disadvantages of implementing this metric on the tool. Such responses may explain why a substantial portion of journal tool metrics that were discussed during focus groups were identified as ‘neutral’ on the 9-point Likert scale [34] within survey responses. Scholarly communication literacy has been identified as essential in the identification of suboptimal transparency practices [35–37]. Increasing literacy through resources, awareness, and education (e.g., workshops) is recommended to combat such practices and maintain research integrity [35,38–40]. Most survey participants were either in mid-level (29.8%, n = 183) or senior (50%, n = 307) career stages. Consequently, there is likely a need to enhance literacy not only for new researchers and clinicians [38,41], but also for individuals at all career stages. For the JTT, participants suggested adding descriptions to explain the nature and purpose of metric items to allow users, especially those who may not be familiar with certain metrics, to make informed decisions when selecting journals. This recommendation is likely going to be implemented on the tool.

Similar to the findings of our earlier study conducted on patient preferences for a JTT [15], it is clear that the tool is unlikely to meet *all* the expressed preferences of participants. However, following the survey and, if not, during focus group voting, a general agreement threshold of above 70% was able to be met for most items. Participants showed support for the following: the implementation of a website user interface, full tool automation, not paying for access, and the implementation of 17 out of 17 suggested metrics to evaluate journals on the tool. Some support for these items was contradictory in nature. For instance, survey participants supported the use of full automation but believed that metrics (e.g., if the written content presented on the website is clear or not) that cannot be fully automated should be implemented. This may be due to low scholarly information literacy among participants. Nevertheless, these features are likely to be considered and utilized during tool development.

The only item during the survey and focus group voting that did not meet the general threshold of 70% was the use of registration. While having some benefits, registration being contentious due to it being a barrier to access aligns with prior research [42]. Participants provided suggestions to increase the value of registration, including adding additional features to track journal metrics over time, linking the tool with ORCID, and having two types of accounts (i.e., one where users do not need to register and another where users register to access specialized features). If registration is implemented on the tool, these additional features are likely to be utilized to address researcher and clinician preferences.

This study represents one study of a three-part initiative to determine the needs and preferences of stakeholder groups (i.e., patients [15], researchers/clinicians, and publishers [16]) for a JTT. Considering the needs of the researchers and clinicians within this analysis allows for the development of a tool that resonates with the community and improves, or otherwise positively impacts, the ability of users to interact with scholarly journals and their published content. Our tool will not only enable us to spotlight the transparency practices of journals but also to track the evolution of these practices over time.

This study has several strengths and limitations. One strength of the present analysis is that we have a heterogenous sample of survey participants, with respondents working in over 75 countries and originating from a variety of researcher and clinician backgrounds (e.g., professors, students, researchers affiliated with government and industry). Utilizing a mixed-methods study with both a cross-sectional survey and focus groups is another strength. While the survey enabled us to determine general journal preferences, the focus groups allowed us to acquire a greater understanding of the reasons behind *why* participants supported and/or opposed JTT features within the survey. One study weakness is that we only included participants fluent in the English language; thus, our findings may not be representative of individuals who do not publish in English. Furthermore, with 632 participants responding to the survey and 22 participants taking part in the focus groups, this analysis had a modest response rate. However, this is not out of line with other online surveys that have used similar recruitment strategies [23,24]. Our response rate is likely to be an underestimation because some emails may be inactive or invalid due to changes in the author’s profession, retirement, or death. Further, with substantially fewer participants taking part in the focus groups (n = 22) compared to the survey (n = 632), the responses provided within the focus groups may not be representative of participant preferences at large. Lastly, our participant sample may also be subject to non-response bias, a difference in response between respondents and non-responders [43], because researchers and clinicians with an interest in transparency practices and/or the implementation of a JTT are more likely to partake in the survey.

## Conclusion

The aim of this mixed-methods study was to obtain researcher and clinician preferences to inform the user design and technological development of a JTT. This two-part analysis involved a cross-sectional survey of authors from biomedical journals identified through MEDLINE, followed by focus groups that were informed by the survey results. Results support the necessity of this tool and suggest that additional initiatives must be undertaken to increase scholarly communication literacy amongst researchers and clinicians. The findings from this study will contribute to refining the JTT and ensuring that it effectively addresses the requirements of the researcher and clinician communities as a means of increasing transparency and open practices within the scholarly community.

## Data Availability

All relevant study materials and data are included in this manuscript or posted on the Open Science Framework.

https://doi.org/10.17605/OSF.IO/6EWQS

## List of Abbreviations

API: Application Programming Interface
CHERRIES: Checklist for Reporting Results of Internet E-Surveys
COREQ: Consolidated Criteria for Reporting Qualitative Studies
DOAJ: Directory of Open Access Journals
DOI: Digital Object Identifiers
JTT: Journal Transparency Tool
MEDLINE: Medical Literature Analysis and Retrieval System Online
ORCID: Open Researcher and Contributor ID
OSF: Open Science Framework
TOP: Transparency and Openness Promotion

## Declarations

### Ethics Approval and Consent to Participate

Research ethics approval was obtained from the Ottawa Health Science Network Research Ethics Board (REB ID # 20230041-01H). The final protocol was registered using the Open Science Framework (OSF) [17] and can be found at https://doi.org/10.17605/OSF.IO/6EWQS

### Consent for Publication

All authors consent to this manuscript’s publication.

### Availability of Materials and Data

All relevant study materials and data are included in this manuscript or posted on the Open Science Framework: https://doi.org/10.17605/OSF.IO/AS3CY

### Authors’ Contributions

JYN: assisted with the design and conceptualization of the study, collected and analysed data, co-drafted the manuscript, and gave final approval of the version to be published.

HL: collected and analysed data, made critical revisions to the manuscript, and gave final approval of the version to be published.

MM: collected and analysed data, co-drafted the manuscript, and gave final approval of the version to be published.

JK: collected and analysed data, made critical revisions to the manuscript, and gave final approval of the version to be published.

DM: designed and conceptualized the study, assisted with the analysis of data, made critical revisions to the manuscript, and gave final approval of the version to be published.

AE: assisted with the analysis of data, made critical revisions to the manuscript, and gave final approval of the version to be published.

AI: assisted with the analysis of data, made critical revisions to the manuscript, and gave final approval of the version to be published.

KDC: designed and conceptualized the study, assisted with the analysis of data, made critical revisions to the manuscript, and gave final approval of the version to be published.

## References

[1] WMA - The World Medical Association. https://www.wma.net/.

[2] Robishaw JD, DeMets DL, Wood SK, Boiselle PM, Hennekens CH. Establishing and Maintaining Research Integrity at Academic Institutions: Challenges and Opportunities. Am J Med 2020;133:e87–90. 10.1016/j.amjmed.2019.08.036.

[3] Zhaksylyk A, Zimba O, Yessirkepov M, Kocyigit BF. Research Integrity: Where We Are and Where We Are Heading. J Korean Med Sci 2023;38:e405. 10.3346/jkms.2023.38.e405.

[4] Lee CJ, Moher D. Promote scientific integrity via journal peer review data. Science 2017;357:256–7. 10.1126/science.aan4141.

[5] Brezgov S. Google Scholar is Filled with Junk Science 2019. https://scholarlyoa.com/google-scholar-is-filled-with-junk-science/.

[6] Hughes B, Joshi I, Lemonde H, Wareham J. Junior physician’s use of Web 2.0 for information seeking and medical education: A qualitative study. Int J Med Inf 2009;78:645–55. 10.1016/j.ijmedinf.2009.04.008.

[7] Duran-Nelson A, Gladding S, Beattie J, Nixon LJ. Should We Google It? Resource Use by Internal Medicine Residents for Point-of-Care Clinical Decision Making: Acad Med 2013;88:788–94. 10.1097/ACM.0b013e31828ffdb7.

[8] Weng Y, Kuo KN, Yang C, Lo H, Shih Y, Chiu Y. Information-searching behaviors of main and allied health professionals: a nationwide survey in Taiwan. J Eval Clin Pract 2013;19:902–8. 10.1111/j.1365-2753.2012.01871.x.

[9] Boeker M, Vach W, Motschall E. Google Scholar as replacement for systematic literature searches: good relative recall and precision are not enough. BMC Med Res Methodol 2013;13:131. 10.1186/1471-2288-13-131.

[10] Haddaway NR, Collins AM, Coughlin D, Kirk S. The Role of Google Scholar in Evidence Reviews and Its Applicability to Grey Literature Searching. PLOS ONE 2015;10:e0138237. 10.1371/journal.pone.0138237.

[11] Manca A, Moher D, Cugusi L, Dvir Z, Deriu F. How predatory journals leak into PubMed. Can Med Assoc J 2018;190:E1042–5. 10.1503/cmaj.180154.

[12] Journal Transparency Tool. Cent Journalology. http://www.ohri.ca/auditfeedback.

[13] Dopp AR, Parisi KE, Munson SA, Lyon AR. A glossary of user-centered design strategies for implementation experts. Transl Behav Med 2019;9:1057–64. 10.1093/tbm/iby119.

[14] Olson GM, Olson JS. User-centered design of collaboration technology. J Organ Comput 1991;1:61–83. 10.1080/10919399109540150.

[15] Ricketts A, Lalu MM, Proulx L, Halas M, Castillo G, Almoli E, et al. Establishing Patient Perceptions and Preferences for a Journal Authenticator Tool To Support Health Literacy: A Mixed-Methods Survey and Focus Group Study. In Review; 2021. 10.21203/rs.3.rs-875992/v1.

[16] Ng J, Moher D, Lalu M, Aalbersberg IjJ, Alperin J, Willinsky J, et al. Publisher Preferences for a Journal Transparency Tool: A Delphi Study Protocol 2022. 10.17605/OSF.IO/UR67D.

[17] Open Science Framework (OSF). https://osf.io/.

[18] Ng JY, Moher D, Ehrlich A, Iorio A, Wicherts J, Grudniewicz A, et al. Researcher and Clinician Preferences for a Journal Transparency Tool: A Mixed-Methods Survey and Focus Group Study 2023. 10.17605/OSF.IO/AS3CY.

[19] Eysenbach G. Improving the Quality of Web Surveys: The Checklist for Reporting Results of Internet E-Surveys (CHERRIES). J Med Internet Res 2004;6:e34. 10.2196/jmir.6.3.e34.

[20] Tong A, Sainsbury P, Craig J. Consolidated criteria for reporting qualitative research (COREQ): a 32-item checklist for interviews and focus groups. Int J Qual Health Care J Int Soc Qual Health Care 2007;19:349–57. 10.1093/intqhc/mzm042.

[21] SurveyMonkey. https://www.surveymonkey.com/.

[22] Aria M. pubmedR: Gathering Metadata About Publications, Grants, Clinical Trials from “PubMed” Database 2020. https://cran.r-project.org/web/packages/pubmedR/index.html (accessed March 16, 2024).

[23] Willis JV, Ramos J, Cobey KD, Ng JY, Khan H, Albert MA, et al. Knowledge and motivations of training in peer review: An international cross-sectional survey. PLOS ONE 2023;18:e0287660. 10.1371/journal.pone.0287660.

[24] Cobey KD, Monfaredi Z, Poole E, Proulx L, Fergusson D, Moher D. Editors-in-chief perceptions of patients as (co) authors on publications and the acceptability of ICMJE authorship criteria: a cross-sectional survey. Res Involv Engagem 2021;7:39. 10.1186/s40900-021-00290-1.

[25] Guest G, Namey E, McKenna K. How Many Focus Groups Are Enough? Building an Evidence Base for Nonprobability Sample Sizes. Field Methods 2017;29:3–22. 10.1177/1525822X16639015.

[26] Joffe H, Yardley L. Research Methods for Clinical and Health Psychology. Content Themat Anal 2003:56–68.

[27] Mullen R, Kydd A, Fleming A, McMillan L. A practical guide to the systematic application of nominal group technique. Nurse Res 2021;29:14–20. 10.7748/nr.2021.e1777.

[28] Hsieh H-F, Shannon SE. Three approaches to qualitative content analysis. Qual Health Res 2005;15:1277–88. 10.1177/1049732305276687.

[29] Hsu C-C, Sandford B. The Delphi Technique: Making Sense Of Consensus. Pract Assess Res Eval 2007;12.

[30] Krueger RA. Focus Groups: A Practical Guide for Applied Research. SAGE Publications; 2014.

[31] Rice DB, Skidmore B, Cobey KD. Dealing with predatory journal articles captured in systematic reviews. Syst Rev 2021;10:175. 10.1186/s13643-021-01733-2.

[32] Battineni G, Baldoni S, Chintalapudi N, Sagaro GG, Pallotta G, Nittari G, et al. Factors affecting the quality and reliability of online health information. Digit Health 2020;6:2055207620948996. 10.1177/2055207620948996.

[33] Swanberg SM, Thielen J, Bulgarelli N. Faculty knowledge and attitudes regarding predatory open access journals: a needs assessment study. J Med Libr Assoc JMLA 2020;108:208–18. 10.5195/jmla.2020.849.

[34] Jebb AT, Ng V, Tay L. A Review of Key Likert Scale Development Advances: 1995– 2019. Front Psychol 2021;12.

[35] Ebadi GZ Saman. Promoting Awareness, Reflection, and Dialogue to Deter Students’ Predatory Publishing. Predatory Pract. Sch. Publ. Knowl. Shar., Routledge; 2023.

[36] Dale J, Craft AR. Professional Applications of Information Literacy: Helping Researchers Learn to Evaluate Journal Quality. Ser Rev 2021;47:129–35. 10.1080/00987913.2021.1964337.

[37] Otike F, Bouaamri A, Hajdu Barát Á. Predatory Publishing: A Catalyst of Misinformation and Disinformation Amongst Academicians and Learners in Developing Countries. Ser Libr 2022;83:81–98. 10.1080/0361526X.2022.2078924.

[38] Ciro JB, Pérez JH. Pedagogical strategy for scholarly communication literacy and avoiding deceptive publishing practices. J Librariansh Inf Sci 2023;0. 10.1177/09610006231187686.

[39] Power H. Predatory Publishing: How to Safely Navigate the Waters of Open Access. Can J Nurs Res 2018;50:3–8. 10.1177/0844562117748287.

[40] Teixeira da Silva JA, Moradzadeh M, Adjei KOK, Owusu-Ansah CM, Balehegn M, Faúndez EI, et al. An integrated paradigm shift to deal with ‘predatory publishing.’ J Acad Librariansh 2022;48:102481. 10.1016/j.acalib.2021.102481.

[41] Richtig G, Berger M, Lange-Asschenfeldt B, Aberer W, Richtig E. Problems and challenges of predatory journals. J Eur Acad Dermatol Venereol 2018;32:1441–9. 10.1111/jdv.15039.

[42] Laera E, Gutzman K, Spencer A, Beyer C, Bolore S, Gallagher J, et al. Why are they not accessing it? User barriers to clinical information access. J Med Libr Assoc JMLA 2021;109:126–32. 10.5195/jmla.2021.1051.

[43] Wang X, Cheng Z. Cross-Sectional Studies: Strengths, Weaknesses, and Recommendations. Chest 2020;158:S65–71. 10.1016/j.chest.2020.03.012.

